# Immunochromatographic SARS-CoV-2 IgG antibody assay: a cross-sectional study conducted at Wakayama Medical University in Japan

**DOI:** 10.1101/2021.01.10.21249421

**Authors:** Sadahiro Iwabuchi, Masahiro Katsuda, Yusuke Koizumi, Mayuko Hatai, Mitsue Kojima, Nahomi Tokudome, Shinobu Tamura, Machiko Nishio, Toshikazu Kondo, Masaya Hironisi, Chiemi Kakutani, Hiroki Yamaue, Shinichi Hashimoto

## Abstract

Asymptomatic patients with severe acute respiratory syndrome coronavirus 2 (SARS-CoV-2) infection must be quickly identified and isolated to prevent the spread of the virus. The number of asymptomatic healthy people is completely unknown because they remain untested. Detection of specific SARS-CoV-2 antibodies has been widely accepted as a diagnostic test, and an immunochromatographic test, which is simpler and relatively cheaper than other methods, is becoming the gold standard for identifying healthy people who had been infected with SARS-CoV-2 in the past. In this study, 1,528 volunteers who worked at a particular hospital were subjected to an immunochromatographic IgG test for SARS-CoV-2 to determine the ratio of asymptomatic people. Only 12 volunteers (0.79%) were IgG^+^, with no significant background differences in the sex, age, profession, experiences of working at the emergency department or caring for coronavirus disease 2019 patients. If this IgG^+^ ratio was to be extrapolated to Wakayama city’s population, 2,780 out of 3,54,063 people may be asymptomatic for SARS-CoV-2. The results imply that anyone may get infected with SARS-CoV-2 but remain asymptomatic.

## Introduction

Cases of the novel coronavirus disease 2019 (COVID-19) caused by severe acute respiratory syndrome coronavirus 2 (SARS-CoV-2) have been reported worldwide. A total of 13,852 positive cases and 389 deaths were reported between January 16 and April 29, 2020 (1). In June 2020, the number of COVID-19 patients seemed to reduce in Japan; however, in early July 2020, a sudden spike in SARS-CoV-2-infected healthy people (SCV-IHP) was reported. Until September 24, 2020, approximately 80,386 cases and 1,525 deaths had been reported in Japan, and this was recognized as the second wave of infection.

Some people with confirmed infection may be asymptomatic but can infect others; this feature is different from that reported during the SARS-CoV infection that emerged in 2003. According to a retrospective study conducted in Beijing around Feb 10, 2020, which was the early phase of SARS-CoV-2 infection, 17.6%, 73.3%, 4.2%, and 5.0% of the COVID-19 patients transferred to hospitals for special treatment had severe, mild, non-pneumonia-like, and no symptoms, respectively (2). On April 7, 2020, 1,095 of 1,453 Chinese COVID-19 patients (75.3%) were asymptomatic (3), and approximately 50%–75% of Italian COVID-19 patients were asymptomatic (4). Thus, it is meaningful to investigate the ratio of asymptomatic COVID-19 patients as well as the ratio of SCV-IHP in the city.

Therefore, an immunochromatographic IgG test for SARS-CoV-2 was used. This study was limited to volunteers who worked at Wakayama Medical Hospital or its branch; these volunteers had no symptoms of COVID-19 on the testing day at least. A total of 143,139 or 436 confirmed cases had been reported in Japan or Wakayama Prefecture as of November 30^th^, 2020.

## Material and Methods

### 1. Samples

Volunteers––doctors, nurses, pharmacists, medical technicians, clerks, researchers, and other professionals (childminders, secretaries, medical interns, etc.)––working at Wakayama Medical Hospital or its branch (medical university) having asymptomatic COVID-19 were enrolled. These volunteers sometimes visited the main hospital daily and/or communicated with the main hospital’s staff as the medical university lies adjacent to the main hospital on the same site. The testing days at the main hospital were as follows: 7–9, 14–16, and 30–31 July, 2020. In contrast, the testing days at the branch hospital were 14–15 September, 2020, because some COVID-19 patients were hospitalized until the end of August 2020 in the branch hospital.

This study was approved by the research ethics committee of Wakayama Medical University (approval no.: 2960), and informed consent was obtained from all volunteers by signature to the document.

### 2. Immunochromatographic test

Immunochromatographic IgG tests were performed using a commercially available kit [SARS-CoV-2 antibody detection kit (IgG), RF-NC002, Kurabo Industries Ltd., Neyagawa, Osaka, Japan]. The product accepts serum, plasma, and whole blood as samples and has a sensitivity, specificity, and accuracy of 76.4%, 100%, and 94.2%, respectively. According to the manufacturer’s instructions, 10 μL of whole blood was added to the wells of the test strips, followed by dilution buffer. Blood was collected using a lancet (NIPRO, Kitaku, Osaka, Japan). The IgG response was evaluated after 15 min, 60 min, and 3 h by three researchers. Approximately 80% of the volunteers, who were IgG^+^, were tested again within 2 weeks to verify the false positive rate and positive predictive value. Thirty-two volunteers who were IgG^+^ at the first round of testing were re-evaluated using the same kit, 22 of which were confirmed false positives. The percentage of false positives and the positive predictive value were 1.59% and 31.2%, respectively.

### 3. Data analysis

The statistical analysis was performed by the analysis of variance (ANOVA), followed with unpaired Student’s t-test. The 95% confidence interval (CI) and chi-squared test results were analyzed using BellCurve for Excel.

## Results

A total of 1,528 volunteers with no confirmed COVID-19 diagnosis were enrolled (median age was 37 (19–77) years, and average age was 40.5 ± 12.0 (mean ± S.D.) years). No significant differences were observed in the volunteers’ sex and age. The number of IgG^+^ volunteers in the main and branch hospital was 12 and 0, respectively (Table 1), and the ratio of IgG^+^ volunteers was 0.79% (95% CI: 0.34%–1.22%). Of the 12 IgG^+^ volunteers in the main hospital, 4 were males and 8 were females, and their median age was 36 (34–61) and 38.5 (24–60) years, respectively. The average age of the IgG^+^ volunteers was not significantly different from that of the IgG^-^ ones in total (*p*= 0.36). Of the IgG^+^ volunteers, 3.3% were pharmacists, 1.4% were clerks, 0.6% were nurses, and 0.8% were other professionals; further, there was no significant difference in the incidence of IgG^+^ results based on the volunteers’ professions (*p* = 0.42).

**Table. 1.** Summary of COVID-19 IgG test in *Wakayama Medical University*

|  | Total | Main hospital | Branch hospital |
| --- | --- | --- | --- |
| *# of volunteers* | 1528 | 1379 | 149 |
| *Sex, male (%) / female (%)* | 486 (31.8%) / 1042 (68.1%) | 434 (31.5%) / 945 (68.5%) | 52 (34.8%) / 97 (65.1%) |
| *Age, male / female, Mean ± S.D.<sup>a</sup>* | 40.5 ± 12.0 / 38.2 ± 11.2 | 40.3 ± 11.9 / 37.7 ± 11.1 | 42.7 ± 13.0 / 42.7 ± 11.5 |
| *Median (range)* | 38 (20–77) / 37 (19–64) | 38 (20–77) / 36 (19–64) | 39.5 (23–68) / 43 (22–64) |
| *Male; Doc, Nur, Pha, Med, Cle, Res, Oth* | 174, 67, 14, 66, 101, 44, 20 | 158, 62, 13, 47, 91, 44, 19 | 16, 5, 1, 19, 10, 0, 1 |
| *IgG positive* | 0, 2, 0, 0, 1, 0, 1 | 0, 2, 0, 0, 1, 0, 1 | 0, 0, 0, 0, 0, 0, 0 |
| *% in each profession<sup>b</sup>* | 0, 3.0, 0, 0, 1.0, 0, 5.0 | 0, 3.2, 0, 0, 1.1, 0, 5.3 | 0, 0, 0, 0, 0, 0, 0 |
| *Female; Doc, Nur, Pha, Med, Cle, Res, Oth* | 74, 533, 31, 42, 215, 24, 123 | 72, 468, 28, 30, 205, 24, 118 | 2, 65, 3, 12, 10, 0, 5 |
| *IgG positive* | 0, 3, 1, 0, 3, 0, 1 | 0, 3, 1, 0, 3, 0, 1 | 0, 0, 0, 0, 0, 0, 0 |
| *% in each profession* | 0, 0.6, 3.3, 0, 1.4, 0, 0.8 | 0, 0.6, 3.6, 0, 1.5, 0, 0.9 | 0, 0, 0, 0, 0, 0, 0 |
| *% of IgG positive in total* |  | 0.79% (95%CI; 0.34–1.22%) |  |
| *% of IgG negative in total* |  | 99.21% (95%CI; 98.8–99.7%) |  |

a) The mean age with standard deviation (mean ± S.D.) and median age of male and female volunteers are indicated.
b) Percentage of volunteers according to their profession; the ratio of IgG<sup>+</sup> volunteers among the same profession.
Abbreviation: Doc (Doctor), Nur (Nurse), Pha (Pharmacist), Med (Medical technicians), Cle (Clerk), Res (Researcher), Oth (Others)

A more detailed analysis of the differences in results based on the locations was performed. In the main hospital, only one patient with COVID-19 symptoms had been hospitalized until the end of July 2020, and no hospitalization was recorded on each testing day. The IgG^+^ rate was 0.87% in total (12/1379 cases); 5 of 530 nurses, 1 of 41 pharmacists, 4 of 296 clerks, and 2 of 209 other professionals were IgG^+^. Two out of five nurses in the medical university had experience working in the emergency department, and one nurse also had experience treating COVID-19 patients; in contrast, the number of nurses that had experience working in the emergency department or treating COVID-19 patients in the main hospital was 60 and 27, respectively. Thus, every 1 of 27 nurses (3.7%) who had cared for COVID-19 patients was IgG^+^. The number of doctors or medical technicians who had cared for COVID-19 patients was 15 and 3, respectively, but none were IgG^+^.

Forty-two patients were hospitalized at the branch hospital until September 15, 2020, and one patient was hospitalized and treated even on the day of the inspection (September 14–15, 2020). The number of doctors, nurses, medical technicians, clerks, and other professionals who had cared for COVID-19 patients was 9, 39, 26, 2, and 1, respectively.

## Discussion

Real-time polymerase chain reaction (RT-PCR) for SARS-CoV-2 is the primary tool for detecting the infection; however, the number of inspections was limited during the first wave of infection because of the expensive equipment and limited well-trained technicians. Improvements in the inspection systems contributed to increased RT-PCR coverage, and an increased number of asymptomatic cases and a decreased number of severe COVID-19 cases was reported during the recent second wave of infection. Whether the virus itself has mutated such that it no more induces severe symptoms of pneumonia or whether the increased RT-PCR coverage has exposed individuals with SARS-CoV-2 who did not have overt symptoms remains unknown.

Apart from RT-PCR, detecting specific antibodies for SARS-CoV-2 (IgM or IgG) is considered reliable. Some assays to detect SARS-CoV-2 IgM/IgG antibodies are enzyme-linked immunosorbent assay (5), quantum dot immunofluorescence assay (6), chemiluminescence immunoassay (7), electrochemiluminescence immunoassay (8) and immunochromatographic test (9-11). IgM antibodies are induced upon recent exposure to SARS-CoV-2, whereas IgG antibodies are produced a few days after exposure; however, the induction period is variable among patients, with differences in symptom severity (6). The sensitivity and specificity of IgG detection depend on the methods and samples. For instance, the sensitivity and specificity are 96.8% and 99.8% using a serum sample and a chemiluminescence immunoassay (7), 100% and 100% using a serum sample and an electrochemiluminescence immunoassay (8), 86.1-94.4% and 100% using a serum sample or 88.66% and 90.63% using whole blood samples and an immunochromatographic test, respectively (11,12). In this case, using whole blood samples and the SARS-CoV-2 antibody detection kit (IgG), the sensitivity was 98.4% based on the calculation used (12). The sensitivity may be relatively higher in this examination because of following reasons: 1) the IgG^+^ responses were checked at least twice by > two researchers at 15-min intervals and 2) approximately 78% of cases that were SARS-CoV-2 IgG^+^ were tested again to avoid false-positive results, as described in the Methods section. The specificity might be 100% because the IgG^+^ strips of the kits were checked using whole blood samples from two volunteers who had symptoms of COVID-19.

A similar evaluation using the same SARS-CoV-2 antibody detection kit was performed among patients who visited Kobe City Medical Center General Hospital without COVID-19 symptoms (13). Thirty-three of 1,000 serum samples were IgG^+^ (3.3%; 95% CI: 2.3%–4.6%), and 50,123 people (95% CI: 34,934–69,868) in Kobe city were found to be SCV-IHP. The present study was limited to the volunteers who worked in the hospital and had more chances of close contact with COVID-19 patients. However, the total ratio of IgG^+^ SCV-IHP was 0.79% (95% CI: 0.34%–1.22%), which was lower than that reported in previous studies. The estimated number of SCV-IHP in Wakayama may be 2,780 (95% CI: 2,677–2,883) out of 354,063, based on the results from Wakayama Medical University. Until November 30^th^ 2020, the total number of COVID-19 positive cases confirmed by RT-PCR in Wakayama city was 146, and all were severe/mild symptomatic or asymptomatic cases. If the current study’s results were to be extrapolated to Wakayama city, only 5.25% (146 of 2,780) of the potential COVID-19 positive cases might be figured out based on the estimated IgG^+^ SCV-IHP.

The current findings indicate that there was no bias in the data due to differences in the professions of the IgG^+^ volunteers. The estimated number of IgG^+^ volunteers in the branch hospital who had experience caring for COVID-19 patients was 8–9 based on the calculation. In fact, most of the volunteers in the branch hospital checked whether they had been infected with SARS-CoV-2 by RT-PCR approximately 2 months prior to the IgG testing day, and all of them had tested negative. These results suggest that the hospital staff at Wakayama Medical Hospital and its branch have high awareness regarding infection control at that time.

In conclusion, the immunochromatographic IgG test for SARS-CoV-2 is simple, rapid, and convenient and has a relatively high sensitivity and specificity. In the near future, anyone could test for SARS-CoV-2 infection easily using the immunoglobulin test kit, regardless of them being symptomatic or asymptomatic, leading to prevention of the spread of infection by “super-spreaders.” Further, SCV-IHP need to be identified as they may have very strong IgG antibodies against SARS-CoV-2, which can serve as an artificial antibody to overcome the worldwide pandemic. The study results emphasize that anyone can be infected with SARS-CoV-2 while still being asymptomatic.

## Data Availability

All data referred to in the manuscript.

## Acknowledgements

The authors would like to thank all of volunteers at Wakayama Medical University hospital and Wakayama Medical University Kihoku hospital. We also thank N.Tokudome, M.Hatai, M. Isono and M. Noguchi for the technical and administrative assistance provided. We would like to thank Editage (www.editage.com) for English language edting. This work was supported by JST CREST Grant Number JPMJCR5G3.

